# Benchmarking the Confidence of Large Language Models in Clinical Questions

**DOI:** 10.1101/2024.08.11.24311810

**Authors:** Mahmud Omar, Reem Agbareia, Benjamin S Glicksberg, Girish N Nadkarni, Eyal Klang

## Abstract

**Background and Aim:** The capabilities of large language models (LLMs) to self-assess their own confidence in answering questions in the biomedical realm remain underexplored. This study evaluates the confidence levels of 12 LLMs across five medical specialties to assess their ability to accurately judge their responses.

**Methods:** We used 1,965 multiple-choice questions assessing clinical knowledge from internal medicine, obstetrics and gynecology, psychiatry, pediatrics, and general surgery areas. Models were prompted to provide answers and to also provide their confidence for the correct answer (0–100). The confidence rates and the correlation between accuracy and confidence were analyzed.

**Results:** There was an inverse correlation (r=-0.40, p=0.001) between confidence and accuracy, where worse performing models showed paradoxically higher confidence. For instance, a top performing model, GPT4o had a mean accuracy of 74% with a mean confidence of 63%, compared to a least performant model, Qwen-2-7B, which showed mean accuracy 46% but mean confidence 76%. The mean difference in confidence between correct and incorrect responses was low for all models, ranging from 0.6% to 5.4%, with GPT4o having the highest differentiation of 5.4%.

**Conclusion:** Better performing LLMs show more aligned overall confidence levels. However, even the most accurate models still show minimal variation in confidence between right and wrong answers. This underscores an important limitation in current LLMs’ self-assessment mechanisms, highlighting the need for further research before integration into clinical settings.

## Introduction

With their capacity to understand and generate human-like text, Large Language Models (LLMs) are poised to support healthcare professionals in complex clinical decisions (1,2). A wide array of LLMs is now accessible, including open source models, offering solutions that cater to both the public and medical professionals (1). The efficacy of these models has been demonstrated in a variety of tasks, albeit with some limitations (3,4). For instance, LLMs like Generative Pre-trained Transformers (GPT) have shown promise in providing diagnostic assistance and answering medical queries (3,5–7). Katz et al. demonstrated that GPT-4 not only improved clinically compared to its predecessor GPT-3.5, but also matched physician performance in certain areas (8). However, there is evidence of “hallucinations” and inaccuracies in model outputs, which could lead to harm in clinical decision-making (9,10). Specifically, LLMs occasionally generated completely fabricated evidence (e.g., information and references) and presented it as factual (9,10).

One way of building confidence in applying models in health care is explainable AI (11,12). However, easily explainable outputs are difficult due to the complexity of how LLMs process and output data (11,13,14). Recent studies show that LLMs tend to be overly confident (15), and it is hard to ascertain where that confidence stems from in a black box system.

The goal of the current study was to benchmark widely used LLMs in terms of accuracy and associated confidence in answering clinical questions. Our aim was to determine if these models can accurately judge when to be confident in their responses, and in doing so, allow for better explainability in their application.

## Materials and Methods

### Study Design and Data Source

This study utilizes a public compiled dataset from a previous study by Katz et al., which includes 655 questions across five medical specialties: internal medicine, obstetrics and gynecology (OBGYN), psychiatry, pediatrics, and general surgery (8). Crafted from internationally recognized textbooks and guidelines, this dataset serves as a standardized framework for assessment (16–20). To enhance benchmarking reliability, each original question was rephrased twice using GPT-4 Application Programming Interface (API) in Python, yielding 1965 questions. The prompts ensured preservation of the original medical content and correct answers (21). A random 20% sample from each field underwent manual validation by two expert physicians, confirming accuracy and consistency with the original questions.

### Model Setup and Configuration

The LLMs employed in this study were prompted to return the correct answer along with a confidence score for each choice (“A”, “B”, “C”, “D”), expressed as a percentage. The open access models were executed using API codes in a dedicated server with 4xH100 80GB GPUs, with the corresponding codebase accessible on GitHub. We utilized Python (3.10) for data analyses. The commercial models were used using the companies API interfaces. We used several Python libraries to facilitate data processing, model interaction, and analysis: NumPy (1.26.4), Pandas (2.1.4, Scikit-Learn (1.3.0), Hugging Face’s Transformers (4.37.2) and torch (2.2.2+cu121), and the Json module (2.0.9). We used the default hyperparameters for each model (22). For the open access models, we used the “instruct” versions, which perform better on zero-shot questioning.

### Benchmarked LLMs

We evaluated 12 LLMs representing a range of model sizes and architectures (**Figure 1**). This selection includes both widely used and recently developed models. The benchmarked models are shown in **Table 1**.

**Figure 1:**
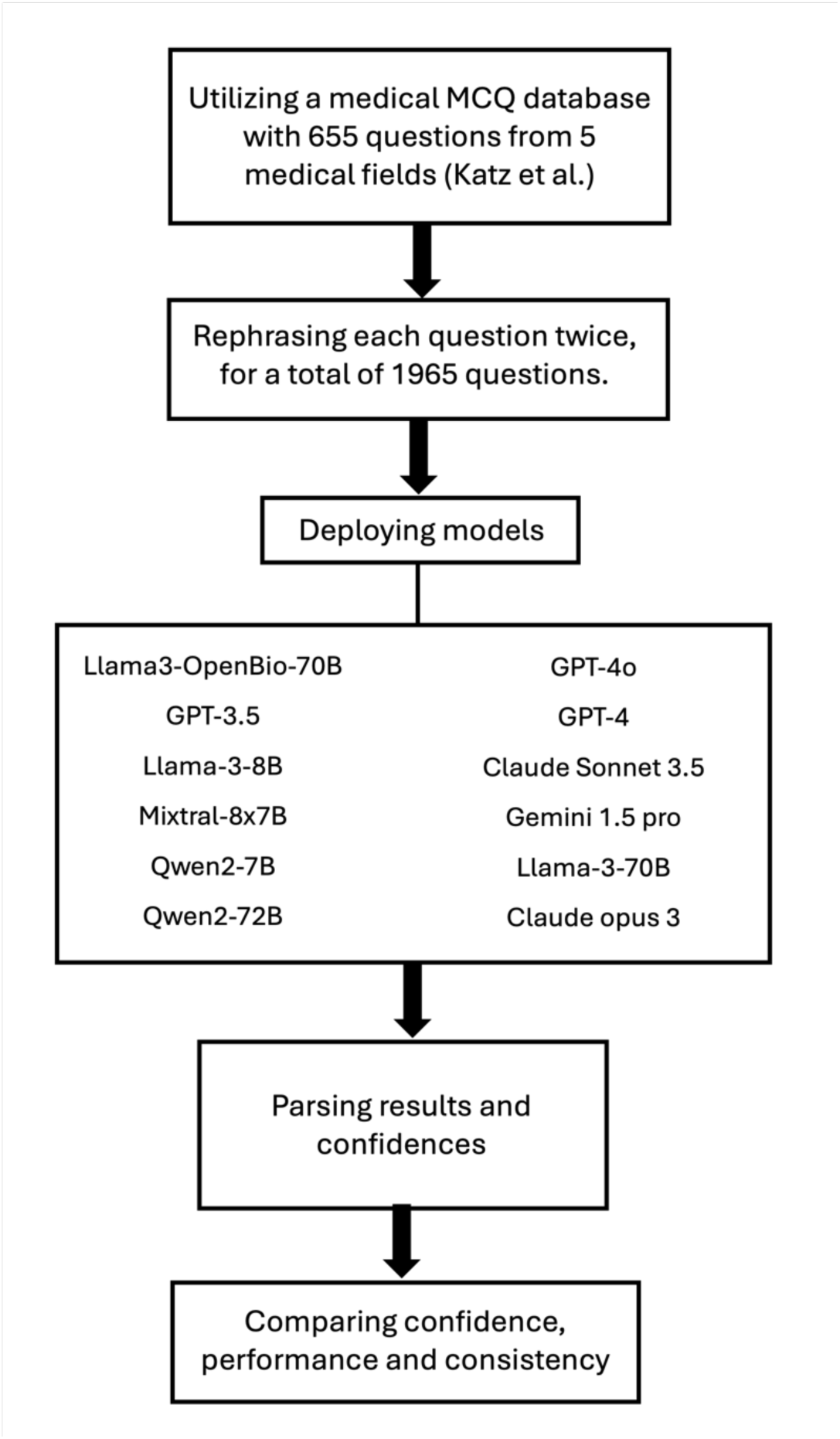
A flowchart representing the evaluation methodology.

**Table 1:**
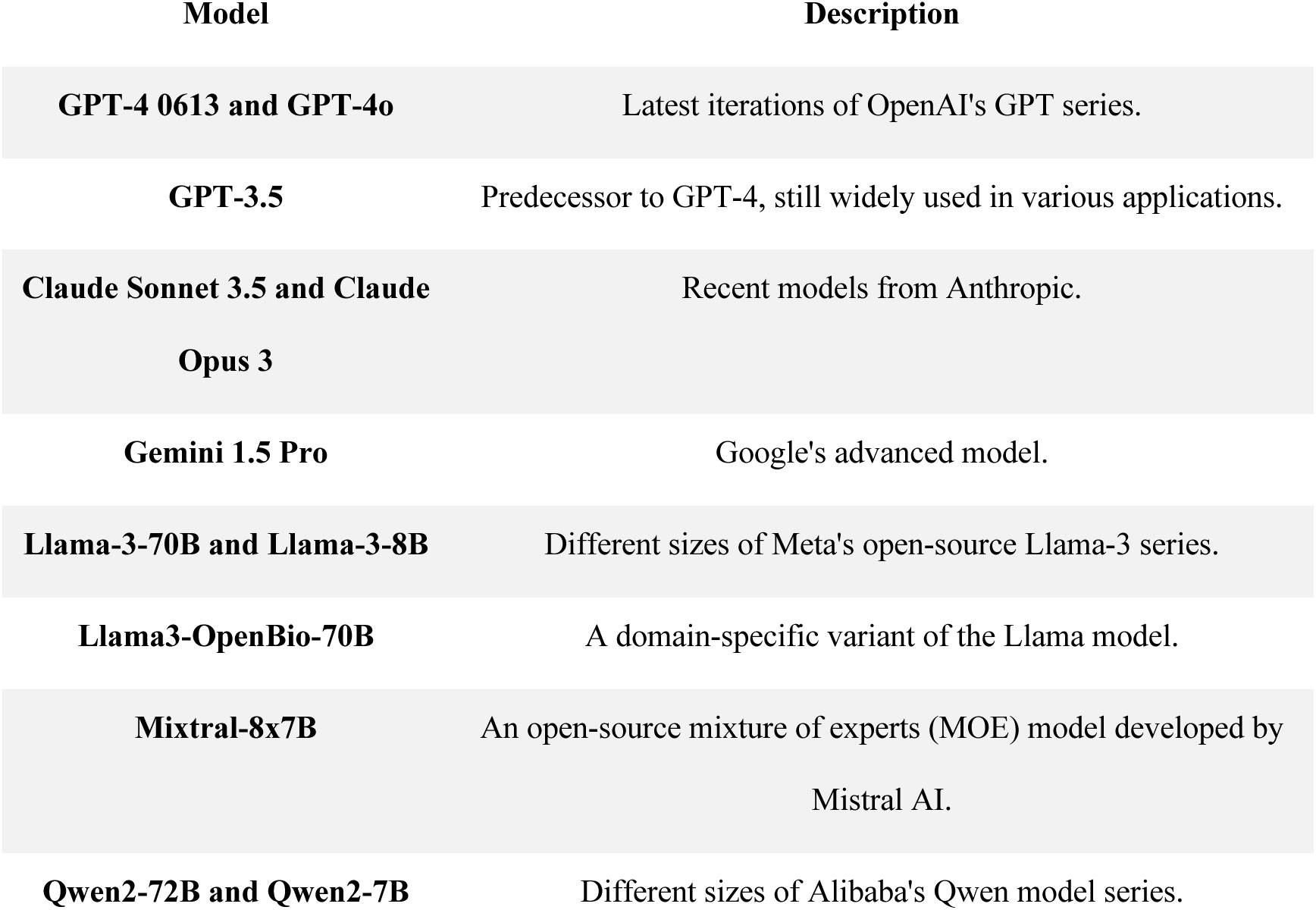
A list of the benchmarked LLMs.

### Statistical analysis

Pearson correlation coefficient was used to correlate means of confidence and accuracy across models and medical fields. Chi-square tests assessed overall performance differences within each field, using proportions of correct responses. Post-hoc pairwise comparisons with Bonferroni correction identified specific inter-model differences. Confidence levels were compared between correct and incorrect responses for each model using two-sample t-tests. Mean confidence scores were calculated for higher-tier and lower-tier models, as well as across all models. Performance consistency was evaluated by comparing confidence gaps between correct and incorrect responses. All statistical tests used a significance level of α = 0.05. Analyses were performed using R version 4.1.2 (R Core Team, 2021).

## Results

### Confidence analysis

**Table 2** summarizes accuracies and confidence levels across the models and **Supplementary Table 1** presents the comprehensive data across all inspected fields and all models. An inverse correlation is demonstrated (r = -0.40, p < 0.01), where better performing models generally show lower confidence.

**Table 2:**
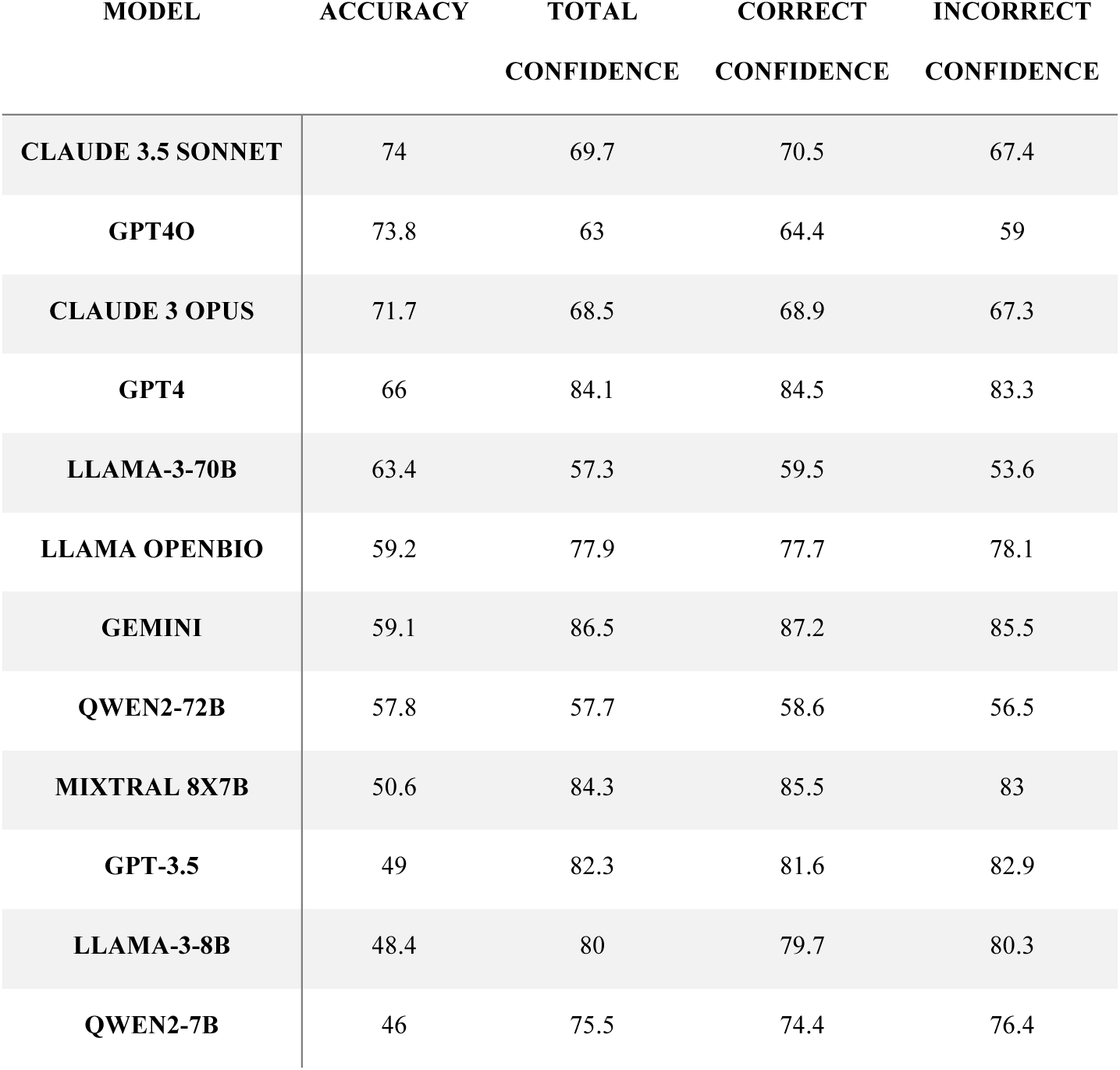
accuracies and confidence levels across the models.

The mean confidence across all 12 models was 76.1% when correct and 74.4% when incorrect. The top six preforming models showed a mean confidence of 72.5% when correct and 69.4% when incorrect, while lower-tier models displayed 79.6% confidence when correct and 79.5% when incorrect. (**Table 3**).

**Table 3:**
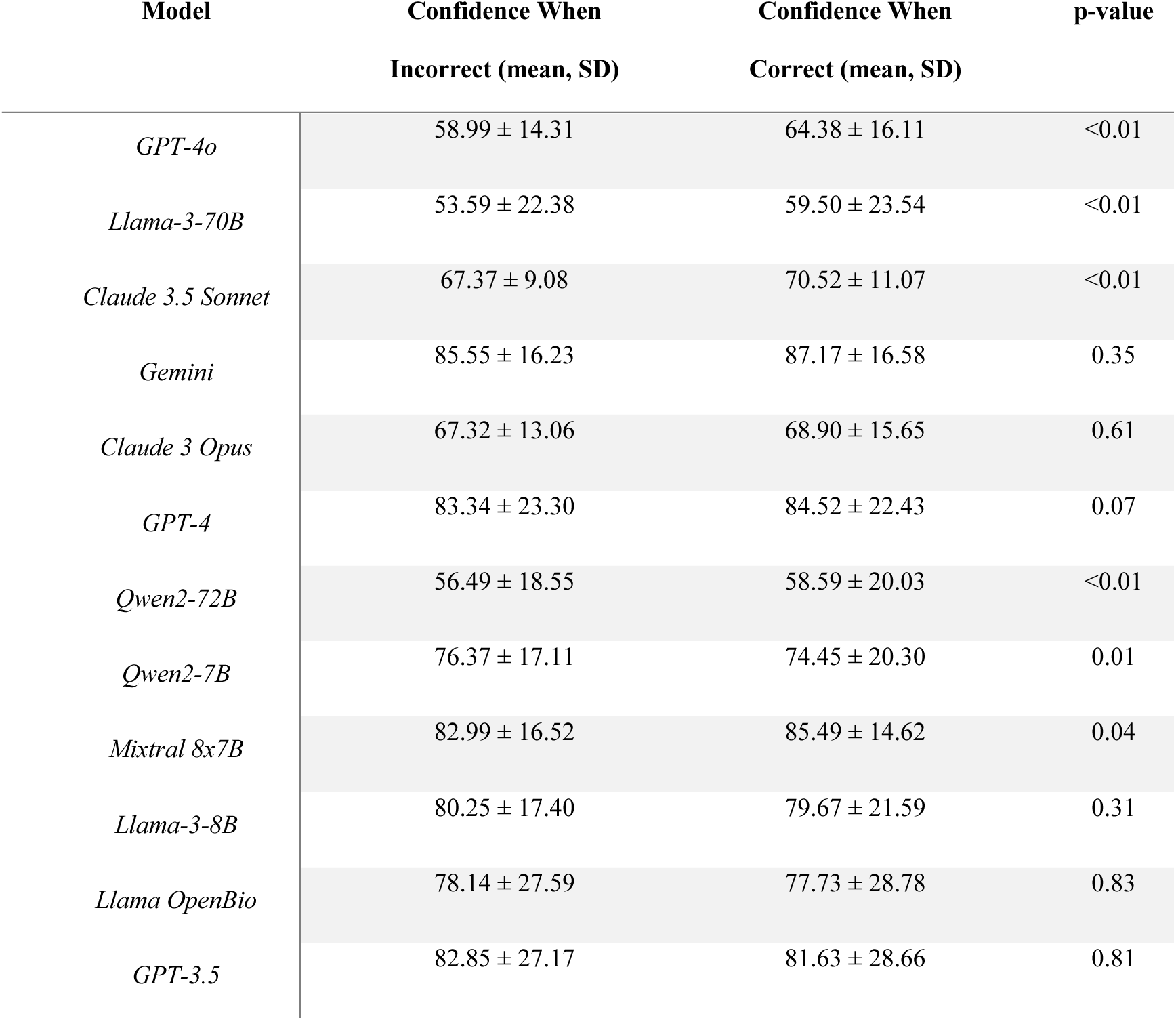
Confidence means of LLMs between correct and incorrect answers.

Four models (GPT-4, Llama-3-70B, Claude 3.5 Sonnet, and Qwen2-72B) demonstrated significantly higher confidence when correct (p < 0.01). Gemini exhibited the highest overall confidence levels (85.6% when incorrect, 87.2% when correct). Qwen2-7B was unique in displaying higher confidence when incorrect (76.4% vs 74.5% when correct, p= 0.01).

GPT-3.5 and Llama-OpenBio-70B revealed minimal differences in confidence between correct and incorrect answers (p = 0.8). The largest confidence gap was observed in GPT-4 (5.4 percentage points), while Llama-3-8B had the smallest gap (0.6 percentage points) (Figure 2).

**Figure 2:**
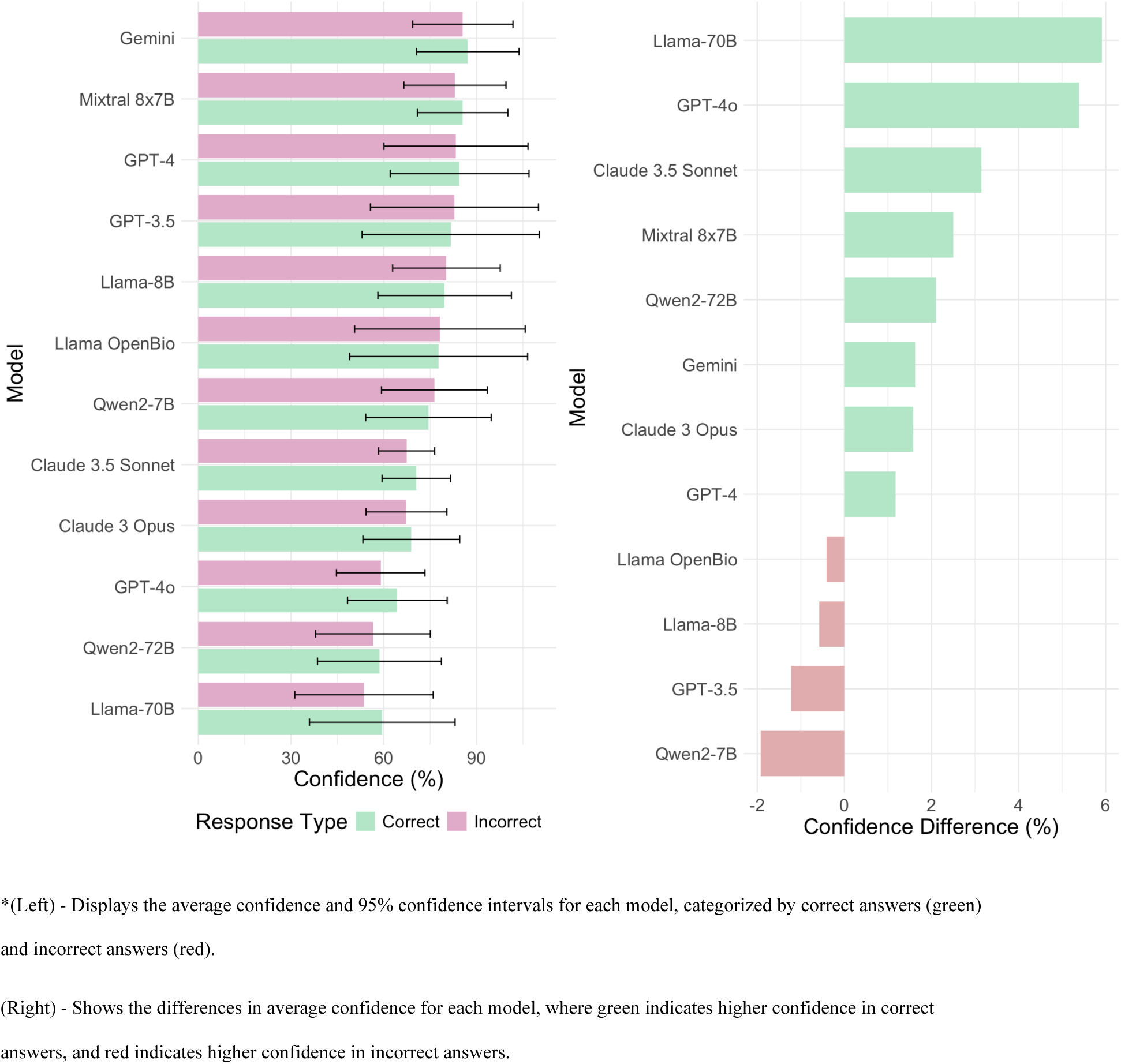
LLMs confidence results between correct and incorrect answers.

### Models’ performances across fields

Significant differences were seen in model performance across all five medical specialties (p < 0.01). GPT-4o and Claude 3.5 Sonnet consistently outperformed other models. In Internal Medicine, GPT-4o (70.9%) and Claude 3.5 Sonnet (73.5%) showed no significant difference (p = 1.0) but outperformed lower-tier models like qwen-7b (43.7%, p < 0.001). For OBGYN, Claude 3.5 Sonnet (71.0%) significantly outperformed most models, including GPT-4 (54.0%, p < 0.001). In Pediatrics, the top five models (GPT-4o, Llama3-70b, Claude 3.5 Sonnet, Claude Opus, GPT-4) showed no significant differences among themselves (all p > 0.05) but outperformed lower-tier models. Psychiatry results mirrored this pattern, with GPT-40 (84.4%) and Claude 3.5 Sonnet (82.4%) leading. In Surgery, GPT-40 (70.9%) and Claude 3.5 Sonnet (70.5%) again showed no significant difference (p = 1.0) but outperformed lower-performing models like qwen-7b (45.6%, p < 0.01) (**Tables 4-5**).

**Table 4:** Top-performing models benchmarking results across different specialties.

| *Specialty* | *Model* | *GPT-4* | *Llama3-70b* | *Claude 3.5 Sonnet* | *Gemini 1.5 Pro* | *Claude 3 Opus* | *GPT-4* |
| --- | --- | --- | --- | --- | --- | --- | --- |
| *Surgery* | Overall | 70.92 | 61.47 | 70.45 | 60.76 | 67.14 | 69.50 |
|  | Original | 70.92 | 55.32 | 66.67 | 58.87 | 65.96 | 68.79 |
|  | Rephrase 1 | 68.79 | 65.25 | 68.79 | 59.57 | 67.38 | 68.79 |
|  | Rephrase 2 | 73.05 | 63.83 | 75.89 | 63.83 | 68.09 | 70.92 |
| *Internal Medicine* | Overall | 75.13 | 58.47 | 73.54 | 58.99 | 70.45 | 64.02 |
|  | Original | 73.81 | 57.14 | 71.43 | 56.35 | 66.67 | 61.11 |
|  | Rephrase 1 | 74.60 | 59.52 | 73.81 | 63.49 | 68.79 | 66.67 |
|  | Rephrase 2 | 76.98 | 58.73 | 75.40 | 57.14 | 75.89 | 64.29 |
| *Obstetrics and Gynecology* | Overall | 65.71 | 55.64 | 70.98 | 58.75 | 65.47 | 53.96 |
|  | Original | 69.06 | 53.96 | 72.66 | 57.55 | 68.35 | 56.12 |
|  | Rephrase 1 | 62.59 | 56.12 | 71.22 | 59.71 | 68.35 | 53.96 |
|  | Rephrase 2 | 65.47 | 56.83 | 69.06 | 58.99 | 59.71 | 51.80 |
| *Pediatrics* | Overall | 71.72 | 68.35 | 71.38 | 57.58 | 68.01 | 67.00 |
|  | Original | 68.69 | 64.65 | 70.71 | 54.55 | 67.68 | 64.65 |
|  | Rephrase 1 | 72.73 | 70.71 | 70.71 | 58.59 | 71.72 | 66.67 |
|  | Rephrase 2 | 73.74 | 69.70 | 72.73 | 59.60 | 64.65 | 69.70 |
| *Psychiatry* | Overall | 84.44 | 73.56 | 82.44 | NA | 79.78 | 77.33 |
|  | Original | 77.33 | 67.33 | 83.33 | NA | 77.33 | 78.00 |
|  | Rephrase 1 | 88.00 | 76.00 | 81.33 | NA | 81.33 | 77.33 |
|  | Rephrase 2 | 88.00 | 77.33 | 82.67 | NA | 80.67 | 76.67 |

**Table 5:** Lower-performing models benchmarking results across different specialties.

| *Specialty* | *Model* | *Qwen-2-72b* | *Qwen-7b* | *Mistral* | *Llama 8b* | *Llama Bio* | *GPT-3.5* |
| --- | --- | --- | --- | --- | --- | --- | --- |
| *Surgery* | Overall | 57.45 | 45.63 | 46.81 | 47.52 | 56.03 | 53.43 |
|  | Original | 53.90 | 46.10 | 45.39 | 46.81 | 58.87 | 52.48 |
|  | Rephrase 1 | 61.70 | 47.52 | 46.10 | 48.23 | 54.61 | 53.90 |
|  | Rephrase 2 | 56.74 | 43.26 | 48.94 | 47.52 | 54.61 | 53.90 |
| *Internal Medicine* | Overall | 47.88 | 43.65 | 44.97 | 40.48 | 53.70 | 45.77 |
|  | Original | 46.03 | 45.24 | 46.83 | 40.48 | 55.56 | 46.83 |
|  | Rephrase 1 | 49.21 | 42.86 | 43.65 | 40.48 | 52.38 | 43.65 |
|  | Rephrase 2 | 48.41 | 42.86 | 44.44 | 40.48 | 53.17 | 46.83 |
| *Obstetrics and Gynecology* | Overall | 55.16 | 43.65 | 44.36 | 43.17 | 53.72 | 42.93 |
|  | Original | 56.83 | 43.17 | 43.88 | 44.60 | 56.83 | 46.04 |
|  | Rephrase 1 | 55.40 | 43.88 | 46.04 | 39.57 | 50.36 | 41.01 |
|  | Rephrase 2 | 53.24 | 43.88 | 43.17 | 45.32 | 53.96 | 41.73 |
| *Pediatrics* | Overall | 59.60 | 39.73 | 52.19 | 48.48 | 61.28 | 49.16 |
|  | Original | 52.53 | 36.36 | 47.47 | 44.44 | 60.61 | 49.49 |
|  | Rephrase 1 | 61.62 | 45.45 | 53.54 | 51.52 | 62.63 | 46.46 |
|  | Rephrase 2 | 64.65 | 37.37 | 55.56 | 49.49 | 60.61 | 51.52 |
| *Psychiatry* | Overall | 67.33 | 54.22 | 63.33 | 60.89 | 70.44 | 57.56 |
|  | Original | 67.33 | 54.00 | 62.00 | 60.00 | 67.33 | 62.67 |
|  | Rephrase 1 | 65.33 | 53.33 | 62.67 | 60.00 | 72.00 | 51.33 |
|  | Rephrase 2 | 69.33 | 55.33 | 65.33 | 62.67 | 72.00 | 58.67 |

## Discussion

In our evaluation, an inverse correlation was demonstrated between accuracy and confidence for LLMs. Notably, some lower-complexity models displayed higher confidence in incorrect answers. However, across all models, there was limited differentiation in confidence levels between right and wrong answers. For instance, GPT-4o, a top performing model, also displayed the largest confidence gap of 5.4 percentage points between correct and incorrect answers. However, this margin is too narrow for practical clinical decision-making. These results highlight potential risks in clinical applications, where model confidence, regardless of answer correctness, could lead to misinformed decisions.

Katz et al. reported GPT-4 outperforming physicians in psychiatry and performing comparably in general surgery and internal medicine (8). Our study corroborates GPT-4’s strong performance, particularly in psychiatry where it achieved 84.4% accuracy. However, our findings suggest more cautious interpretation is needed, given the high confidence levels observed for incorrect answers. Xiong et al.’s work on LLM confidence elicitation aligns with our observations of overconfidence (15). They noted improved calibration and failure prediction as model capability increased, which parallels our finding of better confidence calibration in more complex models. The implications for clinical practice warrant careful consideration. While the performance leap of newer models is promising, their inability to accurately self-assess confidence across wrong answers poses risks. Two possible strategies to address these challenges can be human-in-the-loop protocols and implementing ensemble methods (23).

Human-AI collaboration may offer a balanced approach to leveraging AI strengths while maintaining necessary human oversight in healthcare (24). Sezgin et al. suggested human-in-the-loop approach, ensuring that AI systems are supervised by human expertise. However, effective implementation faces challenges. Careful design of user interfaces is important to prevent automation bias (24,25). There are also concerns about potential erosion of clinical skills with over reliance on AI (26).

Ensemble methods, which aggregate multiple models, present another possible strategy (27). Mahajan et al. conducted review of ensemble learning techniques in disease prediction (28). They found that stacking, an ensemble method that combines multiple classifiers, showed the most accurate performance in 19 out of 23 cases. The voting approach was identified as the second-best ensemble method. However, ensemble methods are computationally intensive and may introduce latency in real-time clinical applications (29).

Both strategies would require extensive clinical trials for validation and the development of model-specific calibration curves for each medical specialty. Our study has several limitations. The dataset was limited to 1965 multiple choice questions across five medical specialties, which may not fully represent the breadth of clinical scenarios. A combination of automatic rephrasing and manual validation could introduce bias (21). Additionally, the study used default model hyperparameters, potentially limiting performance optimization. The generalizability of results to real-world clinical settings remains uncertain. The study did not explore techniques such as fine-tuning and retrieval-augmented generation (RAG), which might improve model accuracy and confidence calibration (30). Finally, the study did not consider the computational cost and time efficiency of deploying these models in practical healthcare scenarios.

In conclusion, better performing LLMs show more aligned overall confidence levels. However, even the most accurate models still show minimal variation in confidence between right and wrong answers. This underscores an important limitation in current LLMs’ self-assessment mechanisms, highlighting the need for further research before integration into clinical settings.

## Data Availability

All data produced in the present study are available upon reasonable request to the authors

## Acknowledgment

We thank Dr. Uriel Katz and the co-authors of the paper “GPT versus Resident Physicians — A Benchmark Based on Official Board Scores, DOI: 10.1056/AIdbp2300192” for sharing the MCQ dataset.

## Competing interest

None declared.

**Ethical approval** was not required for this research, as it utilized already published synthetic data.

---

**Supplementary Table 1.**
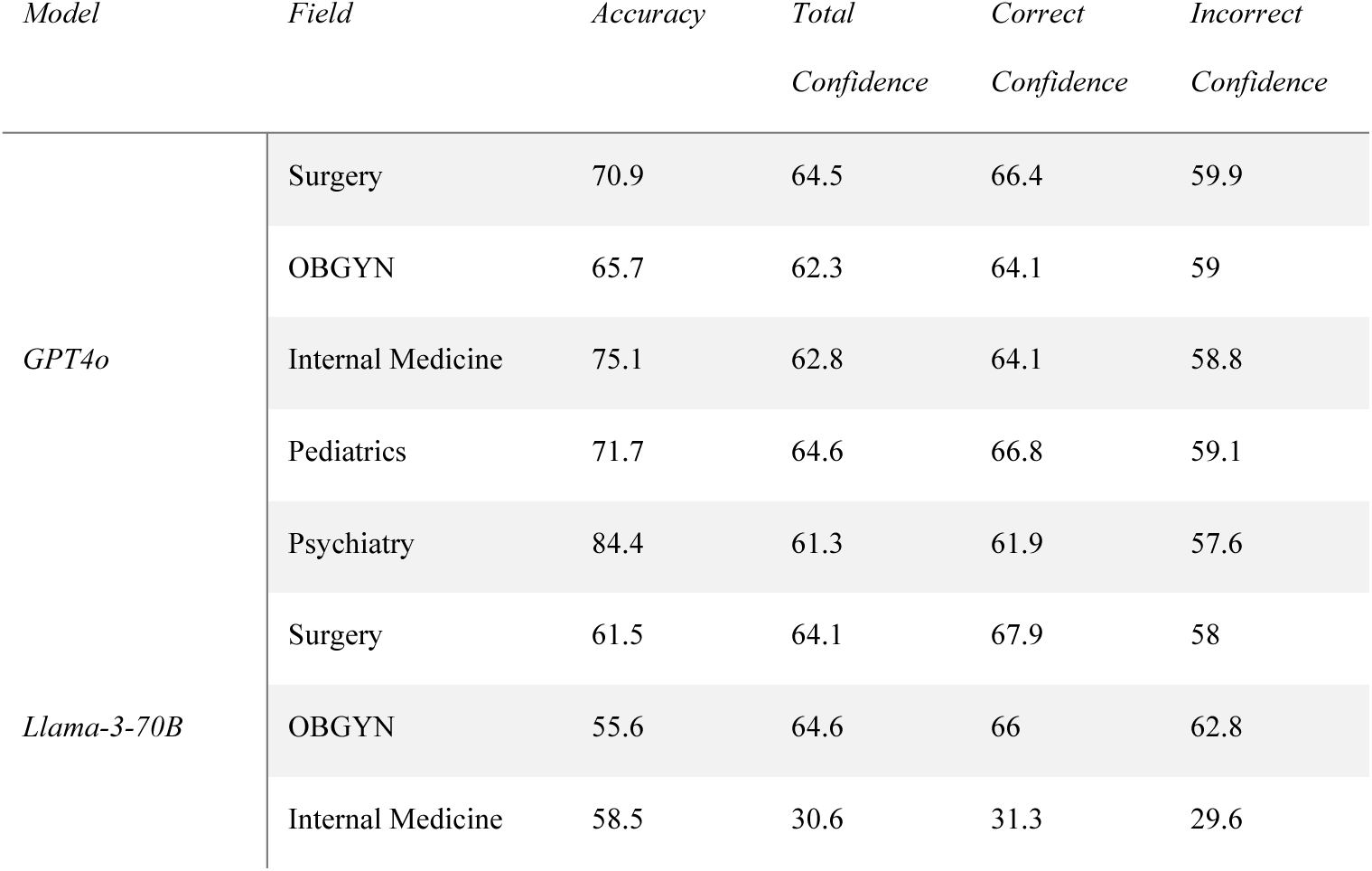
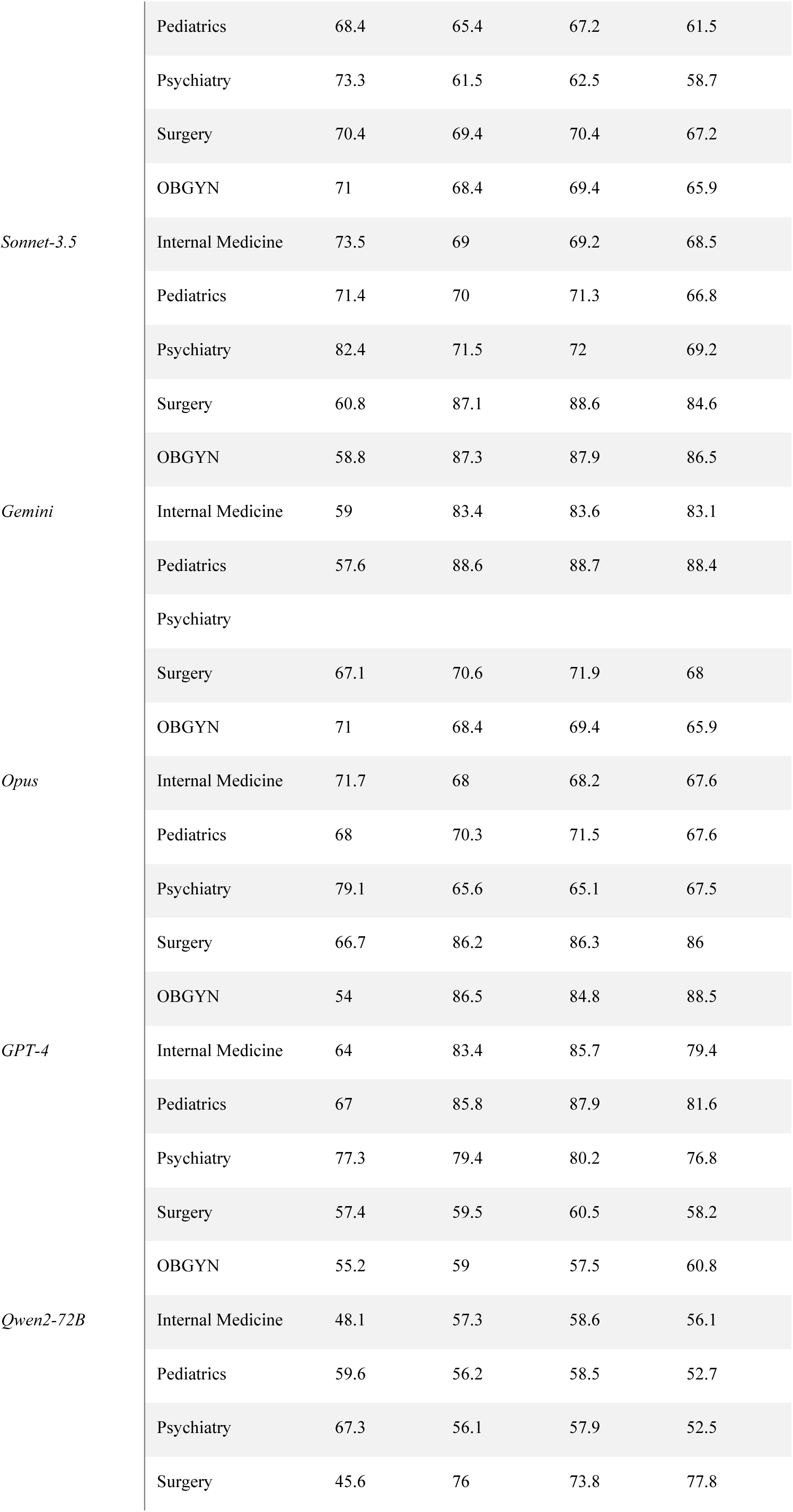
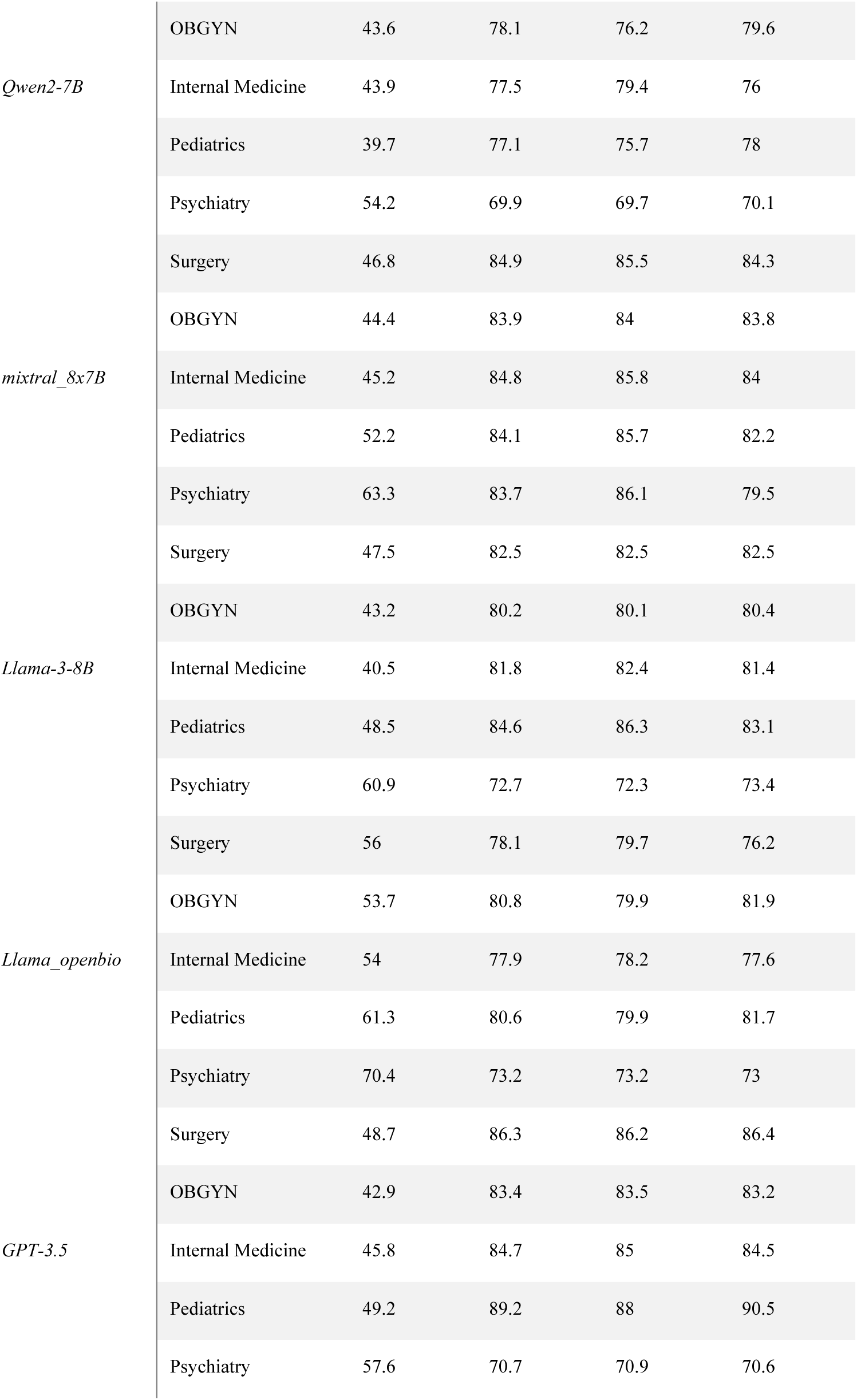

